# Effect of two methods of soft tissue augmentation for socket closure on soft tissue landmarks and ridge dimensions- Results from software-based analysis of clinical data

**DOI:** 10.1101/2022.04.22.22273002

**Authors:** Rampalli Viswa Chandra, Medisetty Bhagyasree, Pulijala Sathwika, Aileni Amarender Reddy

**Affiliations:** Department of Periodontics, SVS Institute of Dental Sciences, Mahabubnagar, Telangana, India

**Keywords:** Periodontics, Alveolar Ridge Augmentation, Tooth Extraction, Gingiva

## Abstract

**Purpose:** The aim of this study was to evaluate the effect of connective tissue grafts (CTG) and pediculated connective tissue grafts (P-CTG) for extraction socket closure on soft tissue landmarks and ridge dimensions.

**Methods:** 45 subjects were randomized into two groups, where the soft tissue closure of extraction sockets was done by pediculated connective tissue grafts (P-CTG group) and connective tissue grafts (CTG group). On photographs taken at different time-points, four markers corresponding to marginal gingiva (MG), mucogingival junction (MGJ), mesial papilla (MP) and distal papilla (DP) were placed. After image alignment and stacking, the markers were tracked through five photographs taken during the course of the therapy. The displacement, total displacement (TD), divergence and angle of maximum divergence (AMD) for each marker were recorded. Buccolingual and mesiodistal dimensions of edentulous sites were measured, and the difference from the baseline to 6-months in both the dimensions was calculated and was described as buccolingual gain (BL gain) and mesiodistal gain (ML gain) respectively.

**Results:** Intergroup comparison of displacement and divergence of markers across both the groups showed varying levels of significance. P-CTG resulted in higher buccolingual dimensions at 3 months (*P*=0.001) and 6 months (*P*=0.002) when compared to the CTG group. From baseline to 6-months, the BL and MD gains were 2.82±1.29mm & 1.68±1.12mm and 1.27±0.65mm & 1.48±0.67mm in P-CTG and CTG group respectively. In the P-CTG group, TD (r=0.4; *P*=0.001) & AMD (r=0.5; *P*=0.05) of MG and TD (r=0.2; *P*=0.005) of MGJ resulted in BL gains. In the CTG group, TD (r=0.7; *P*=0.004) and AMD (r=0.3; *P*=0.004) of MP and AMD of MGJ was associated with a significant BL gain.

**Conclusions:** P-CTG and CTG had significant effects on displacement and divergence of soft tissue landmarks respectively. The relocation of these landmarks, seem to result in an improvement in the buccolingual dimensions of the post-extraction ridge.

## INTRODUCTION

The sequelae of socket collapse and the subsequent hard and soft-tissue remodelling has a significant impact on its aesthetics and function [1-3]. With an expected loss of anywhere between 40 to 60% of the original height and width [3], several socket preservation techniques have been documented to mitigate the effects of inadequate ridge volume and dimensions following tooth extraction [1,3,4].

The dynamic nature of a healing extraction socket frequently leads to change in soft tissue landmarks on the ridge [5,6]. The buccal aspect of the ridge is often the most affected site, [1-3,5,6] resulting in the relocation of the ridge to a more palatal/lingual position [6,7]. Achieving primary closure by advancing the flap over an extraction socket was thought to be necessary [3,4,6,7], however, it may lead to alterations in ridge contours [6], iatrogenic distortion of soft-tissue landmarks such as the muco-gingival junction (MGJ) [8,9] and interdental papilla [6,7] and buccal vestibule [9]. The iatrogenic distortion of soft tissue landmarks can lead to ridge contraction [3,6,7] and loss of attached gingiva, [3] which can impact the aesthetics and function of the site [8,9].

There is now an onus on procedures that limit or reverse this inadvertent damage cause to the ridge; the use of “flapless approach”[2], ‘‘socket-plug’’ techniques [6] and soft tissue grafts [7,8] were advocated primarily to prevent and limit alterations in ridge dimensions and anatomy due to flap advancement alone [3,4,8,9]. Palatal connective tissue grafts (CTG) [10] have been frequently used for correcting ridge deficiencies by virtue of their excellent tissue integration and volume stability [10,11], leading to reduced soft tissue shrinkage and preservation of the gingival architecture. An immediate post-surgical gain of 2 mm is possible [2,4,9,11,12], but because it is a “free” graft, its effectiveness is greatly dependent on its ability to derive blood supply from the recipient bed and the mucoperiosteal flap [7,11,12]. Complete coverage and stability of the graft are crucial and this may necessitate elevation of the flap potentially compromising blood supply to it [7,11,12]. To offset these disadvantages, a pediculated CTG (P-CTG), which maintains its own blood supply has been advocated [13]. A modified technique by El Chaar et al. [14] improves on the conventional technique by using thick donor tissue and by creating a large graft base and a periosteal bed near the extraction socket to promote vascularization. Most importantly, there is minimal elevation of the buccal flap as the graft is only tucked into it and stabilized with sutures, minimizing the displacement and distortion of interdental papillae and the MGJ [13,14] in light of these approaches, the primary hypothesis behind this study was that the P-CTG procedure would result in minimal displacement of gingival landmarks when used for socket grafting.

While the measurement of distances between key anatomical landmarks such as the gingival margin [15], mucogingival junction [11], depth of vestibule [9] and/or peaks of interdental papillae [16] has always been the norm in assessing outcomes, data however is lacking on the dimensional stability of these visible landmarks on the soft tissues after procedures such as soft tissue augmentation for socket closure [11,12]. Existing methods of measurement for soft tissue grafts do not detect the variability of these landmarks during the healing phase nor attempt to interpret the final results in the context of moving landmarks [2,5-7,11,16]; failure to do so might impact the overall and final results. Investigators are also limited by the solutions and tools available at this time [11,17]. However, identifying displacement, at least in two dimensions may provide an insight on their impact on healing and the final result of a procedure.

In this context, the effect of connective tissue grafts (CTG) and pediculated connective tissue grafts (P-CTG) on soft tissue landmarks and ridge dimensions when used for socket closure was investigated. Apart from comparing the clinical effects of the procedures, additional analysis was done on the data based on the principles of image alignment, stacking and tracking.

## MATERIALS AND METHODS

### Sample size calculation

As per proportional power calculation, a minimum sample size of 16 per group (32 in total) was needed to detect a ridge-height difference of 1 mm when the power of the test is 0.80 at a significance level of 0.05.

### Participants and Eligibility Criteria

The initial pool consisted of 122 subjects who reported to our department between January and May, 2019. After scaling and root planing, all subjects underwent a thorough soft-tissue examination including sounding of the tissues. From this pool, 45 subjects satisfying the following inclusion criteria were included in this study; 1. subjects within the age group of 20-50years 2. Subjects requiring single tooth extraction in the anterior area without any signs and symptoms of acute infection or inflammation. 3. Thin periodontium [18] around the affected site characterized with a residual buccal periodontal pocket≥5 mm [19] 4. Subjects with extraction defects type-2 (based on the extraction defect sounding (EDS) [20] classification) and characterized by slight compromise of the local tissues with vertical or transverse soft tissue loss of 1-2 mm and with not more than one compromised socket wall. 5. A presurgical depth of 3 mm in the palatal mucosa of the distal aspect of canine [10,13,14]. 6. Subjects not consenting for hard tissue augmentation/implants because of financial reasons and willing to wear a fiber-reinforced, composite-resin (FRC) bonded temporary bridge with a hygienic pontic for 6-months. 7. Systemically healthy subjects and those who are consenting for single tooth extraction.

### Trial Design and Interventions

A randomized-controlled, single-blind study design was conducted on 45 subjects (Age: 37.87±10.22years; 45 sites;31 males). Double-blinding was not feasible as the nature of surgery and surgical site precludes blinding of operators and investigators. Randomization software [21] was used to randomly allocate subjects into either of the two treatment groups; P-CTG group (test group) and CTG groups (control group) where the soft tissue closure of extraction sockets was done by pediculated connective tissue grafts (P-CTG) and connective tissue grafts (CTG) respectively. Approval from institution ethical committee (SVSIDS/PERIO/1/2017) was obtained and informed consent was taken from all the subjects. All surgeries were performed by two designated operators (RVC & BM) and the readings were recorded by a calibrated set of two investigators (PS & MB).

P-CTG procedure was performed as follows (Figure 1) [14]; flapless atraumatic extraction of the tooth was done to preserve the papillae and interproximal bone. Granulation tissue was removed with curettes and the socket was irrigated with sterile normal saline. The recommended donor site for harvesting the connective tissue graft is from the distal side of the canine to mid-palatal side of the first molar approximately 3-4 mm from the free gingival margin of the hard palate. The sum of (a) buccolingual dimensions of the extraction site (b) 3-4 mm to account for the gap between the extraction site and base of the pedicle and (c) an additional 2-3 mm to facilitate tucking the flap underneath the buccal gingiva of the extraction socket was the length of the pedicle required to close the socket effectively. An epithelial window was raised and the pedicle was elevated. The medial attachment of the P-CTG was maintained. The isthmus between the base of the pedicle and the extraction site was de-epithelialized and free end of the P-CTG was flipped and folded passively over the extraction site and tucked underneath the buccal flap of the extraction socket and secured with 4-0 Prolene sutures (Trulene®, Sutures India, Bengaluru, India). CTG was performed similarly (Figure 2); the buccal and lingual flaps were raised 3 mm beyond the level of crestal bone and the tooth was atraumatically extracted. The classic three-sided “trap door” incision [10] was used to harvest the CTG; The length and breadth of the trapdoor was determined by the dimensions of the graft required. The graft was sutured to the recipient bed with 4-0 Chromic Catgut (TruGut®, Sutures India, Bengaluru, India). Care was taken to ensure that the flaps rested passively on the CTG before suturing. A tension-free approximation of the flap was done with 4-0 Prolene sutures (Trulene®, Sutures India, Bengaluru, India). The baseline and post-operative outcomes were recorded by two calibrated investigators (MB and PS); their mean weighted inter-examiner kappa scores were 0.79, 0.76, 0.89 and 0.76 for image alignment, stacking, bucco-lingual and mesiodistal dimensions respectively. Reproducibility for placement of pre-determined markers was poor (<0.75), hence it was marked only after a verbal agreement between all the three primary investigators of this study (RVC, MB and PS).

**Figure 1:**
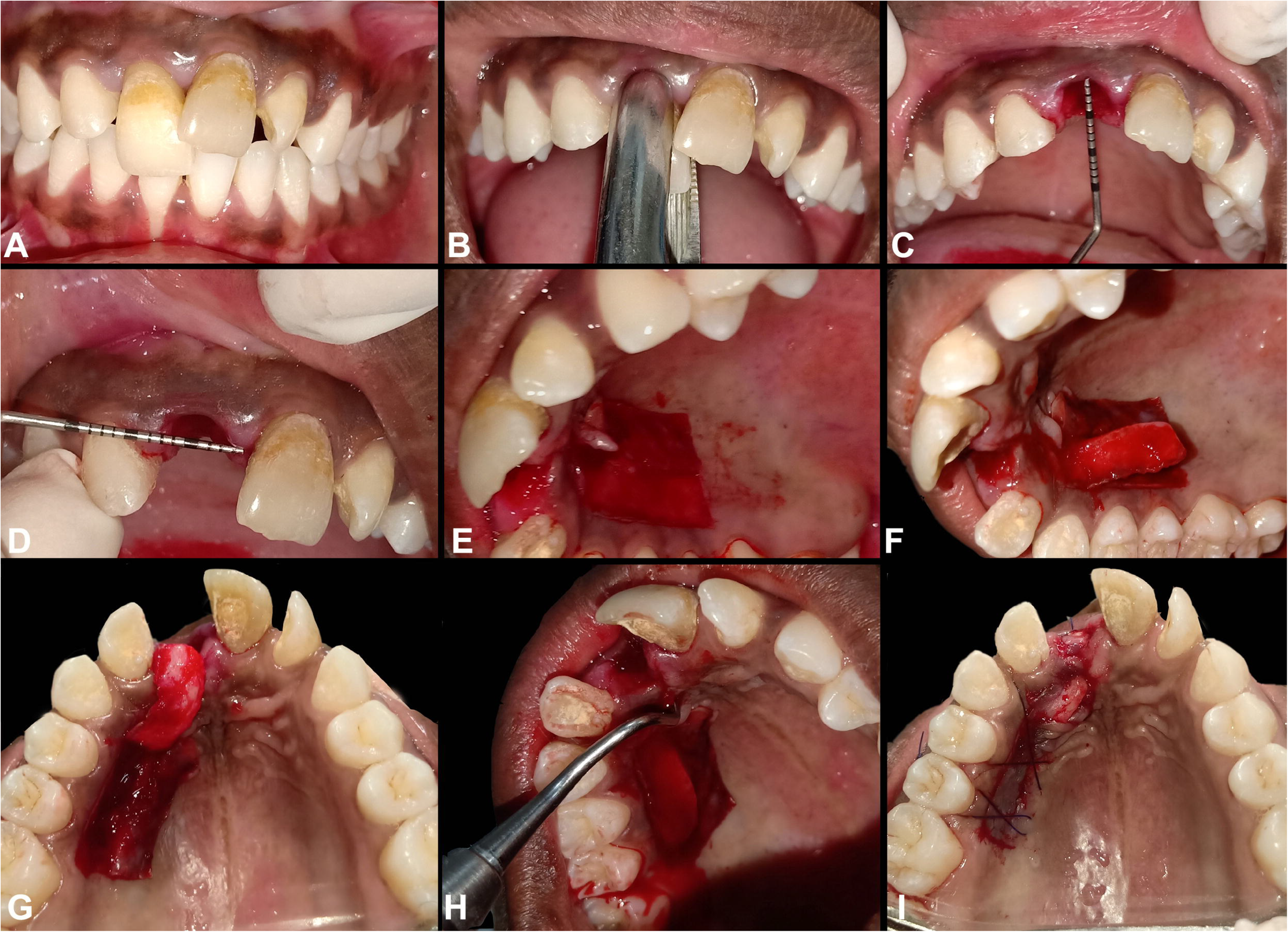
P-CTG Procedure; flapless (A) atraumatic extraction of the tooth (B) was done. The dimensions of the extraction site were measured to determine the length of the pedicle required to close the socket effectively (C; D). An epithelial window (E) was raised and the pedicle was elevated (F). The medial attachment of the P-CTG was maintained (G). The isthmus between the base of the pedicle and the extraction site was de-epithelialized (H) and free end of the P-CTG was tucked underneath the buccal flap of the extraction socket (I).

**Figure 2:**
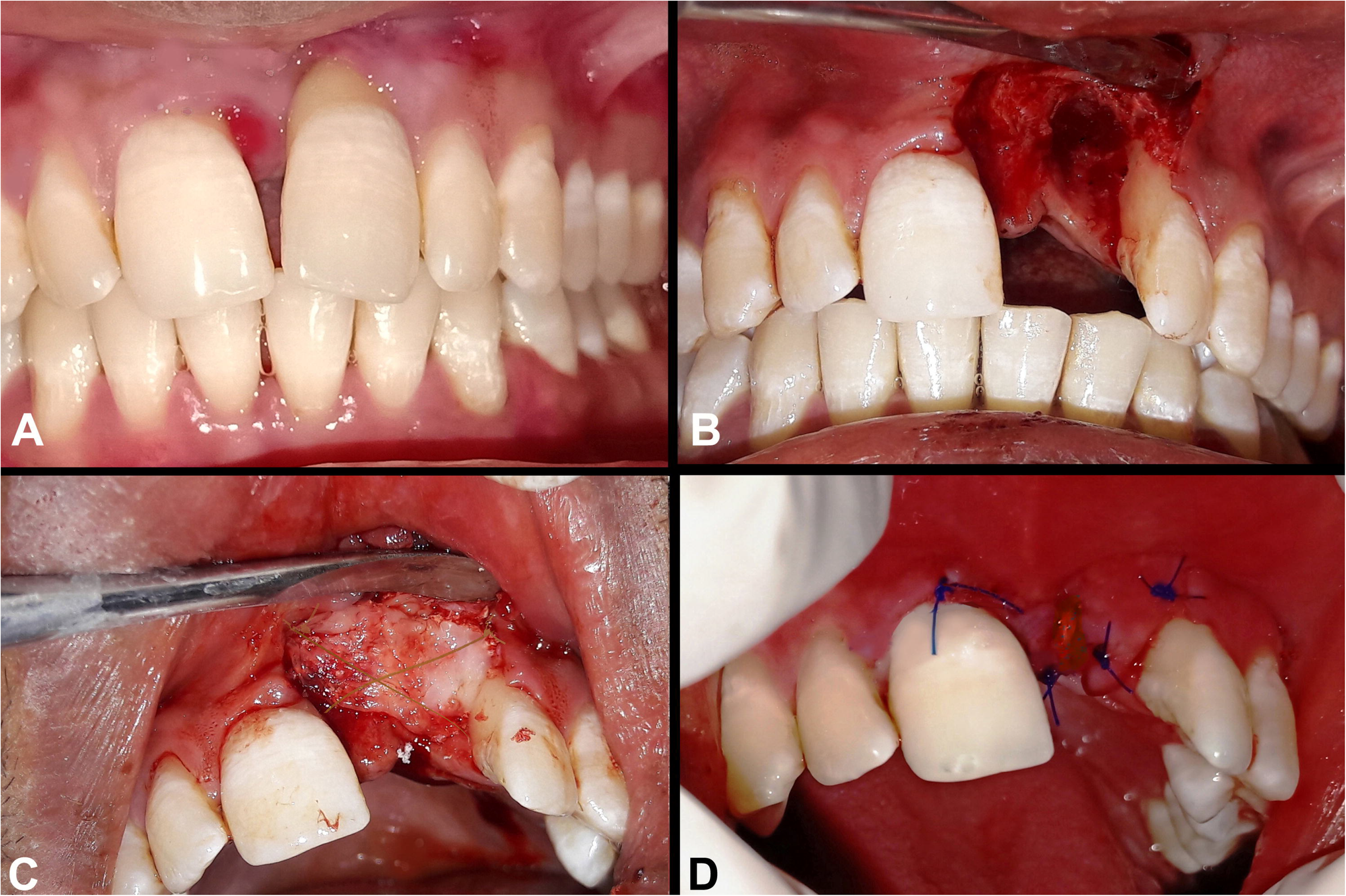
CTG Procedure; after the tooth (A) was atraumatically extracted (B), buccal and lingual flaps were raised 3 mm beyond the level of crestal bone. The harvested CTG was sutured to the recipient bed with 4-0 Chromic Catgut (C). A tension-free approximation of the flap was done with 4-0 Prolene sutures (D).

### Outcomes

#### Image alignment, stacking and tracking of pre-determined markers

At baseline (before the procedure), immediately after surgery (post-OP) and at 1,3 and 6 months, photographs of the subjects in frontal view after adequate retraction with the contact point of the upper central incisors as the center of the image were taken. The protocol of Ricci [22] was adapted to ensure accurate positioning and orientation at all time-intervals. In each photograph, 4 landmarks or “markers” were placed using a photo editing software (Photoshop®, version 21.1.1, Adobe, Noida, India) (Figure 3). The markers were as follows; 1. Marginal Gingiva (MG): The most apical point of the gingival scallop on the marginal gingiva was considered as the marker “MG” 2. Mucogingival Junction (MGJ): As it is a continuous line, the mid-buccal point on the mucogingival junction was considered as the marker “MGJ” 3. Mesial Papilla (MP): The tip of the mesial interdental papilla at its most coronal aspect was considered as “MP” and 4. Distal Papilla (DP): The marker “DP” was the tip of the distal papilla at its most coronal aspect. The marker *(29*26 pixels)* was placed on the brightest pixel at the region-of-interest only after the three investigators (RVC, BM & PS) agreed on its location. After ensuring that all the five pictures (at baseline, post-OP,1, 3 and 6 months) from a single subject were similar in size and orientation, except the markers, the rest of the photograph was darkened by adding a mask of the photograph to itself and then by using the *image>calculations>exclusion* commands in the photo editing software. This reduces the picture “noise” during image stacking, and tracking. A temporal “stack”, which is the set of 5 images related by time [23] was created in the software ImageJ® (LOCI at the University of Wisconsin-Madison, USA). A plugin for the above software, TrackMate® [24] was used to track the movements of these four markers through five photographs. Briefly, this was done as follows [24]: The stack of photographs with the markers was opened in ImageJ® and TrackMate® was launched. The difference of Gaussian detector (DoG detector) was used as a detection algorithm for tracking the markers and after initial and final spot filtering, which help to prevent the software from identifying undesired markers except the ones in question; a linear assignment problem (LAP) tracker was used to track all four markers in the photographs. The data obtained had two components (Figure 4): 1. Displacement and Total Displacement (TD): Displacement is the distance of the marker from one time point to the next subsequent time point and Total displacement (TD) is the distance from baseline position of the marker to its position at 6 months. 2. Divergence and Angle of Maximum Divergence [25]: Divergence is the angle between the expected direction and the observed direction of the marker and angle of maximum divergence is the angle from the source to the final location of the marker. The tracks are composed of four-line segments which correspond to the movement of the trackers between the following time-points: baseline to post-OP, post-OP to 1-month, 1-month to 3-months and 3-months to 6-months (Figure 5).

**Figure 3:**
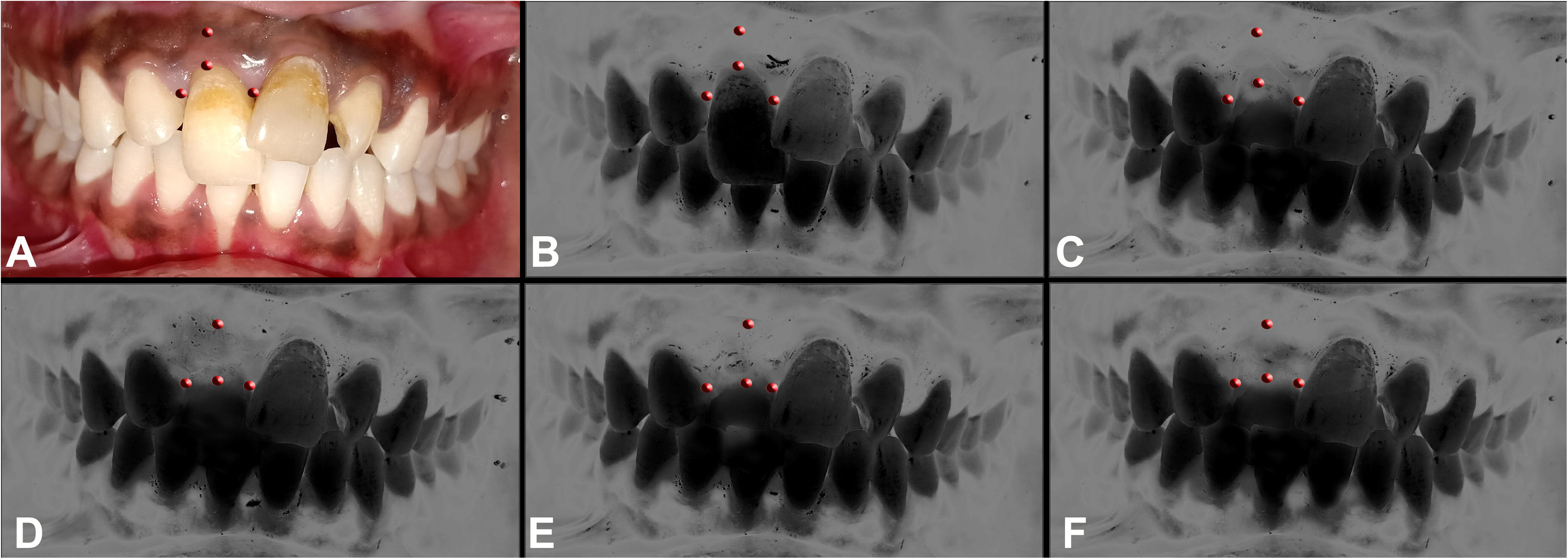
In each photograph, 4 landmarks or “markers” were marked using a photo editing software (A). The markers were 1. Marginal Gingiva (MG) 2. Mucogingival Junction (MGJ) 3. Mesial Papilla (MP) and 4. Distal Papilla (DP). After ensuring that all the five pictures (at Baseline, PostOP,1, 3 and 6 months) from a single subject were similar in size and orientation, except the markers, the rest of the photograph was darkened by adding a mask of the photograph to itself (B to F).

**Figure 4:**
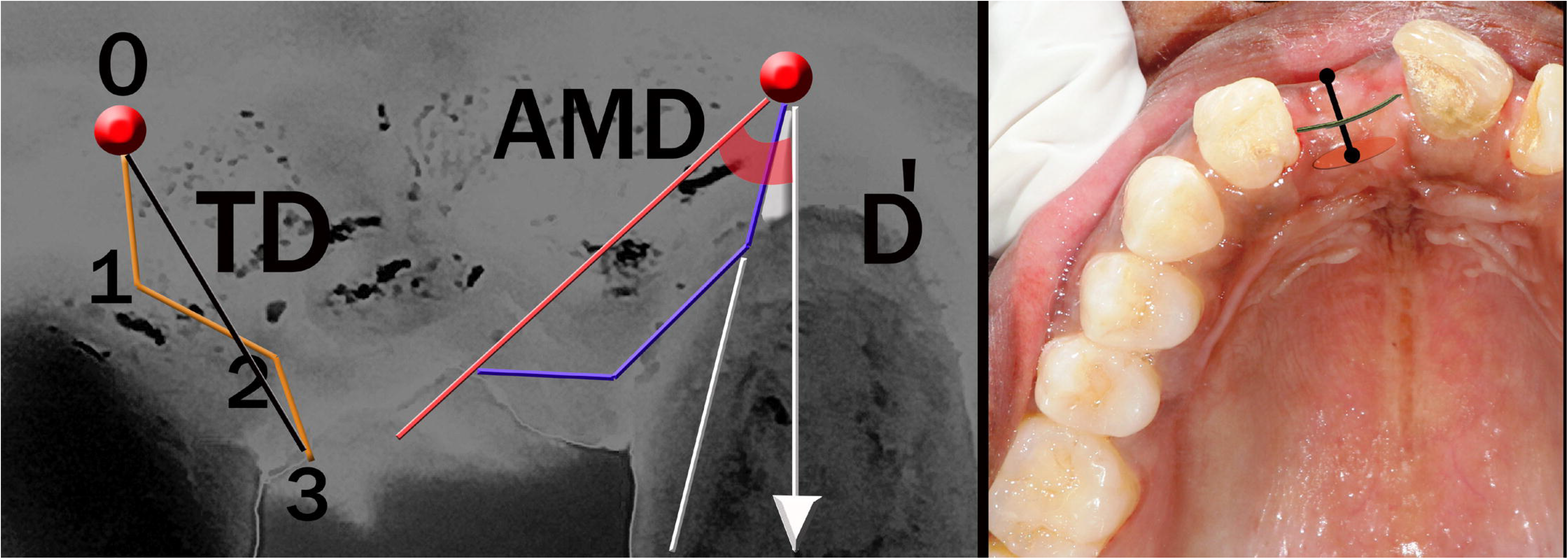
Description of outcomes; Displacement (left panel) is the distance of the marker from one time point to the next subsequent time point (distance from 1 to 2 for example). Total displacement (TD; left panel) is the distance from baseline position of the marker to its final position (distance from 0 to 2). Divergence (D’; left panel) is the angle between the expected direction (arrow) and the observed direction of the marker (white line). Angle of maximum divergence (AMD; left panel) is the angle from the source (arrow) to the final location of the marker (red angle). The distance from the greatest convexity or bulge on the buccal and lingual/palatal soft-tissue beneath the buccal and lingual mucogingival lines or the mid-buccal area immediately beneath the first rugae in the palatal mucosa (middle panel) at the center of the mesio-distal space was considered as the bucco-lingual dimension (right panel). The distance from the crests of the mesial to the distal papilla through the center of the ridge was considered as the mesio distal dimension (middle/right panels).

**Figure 5:**
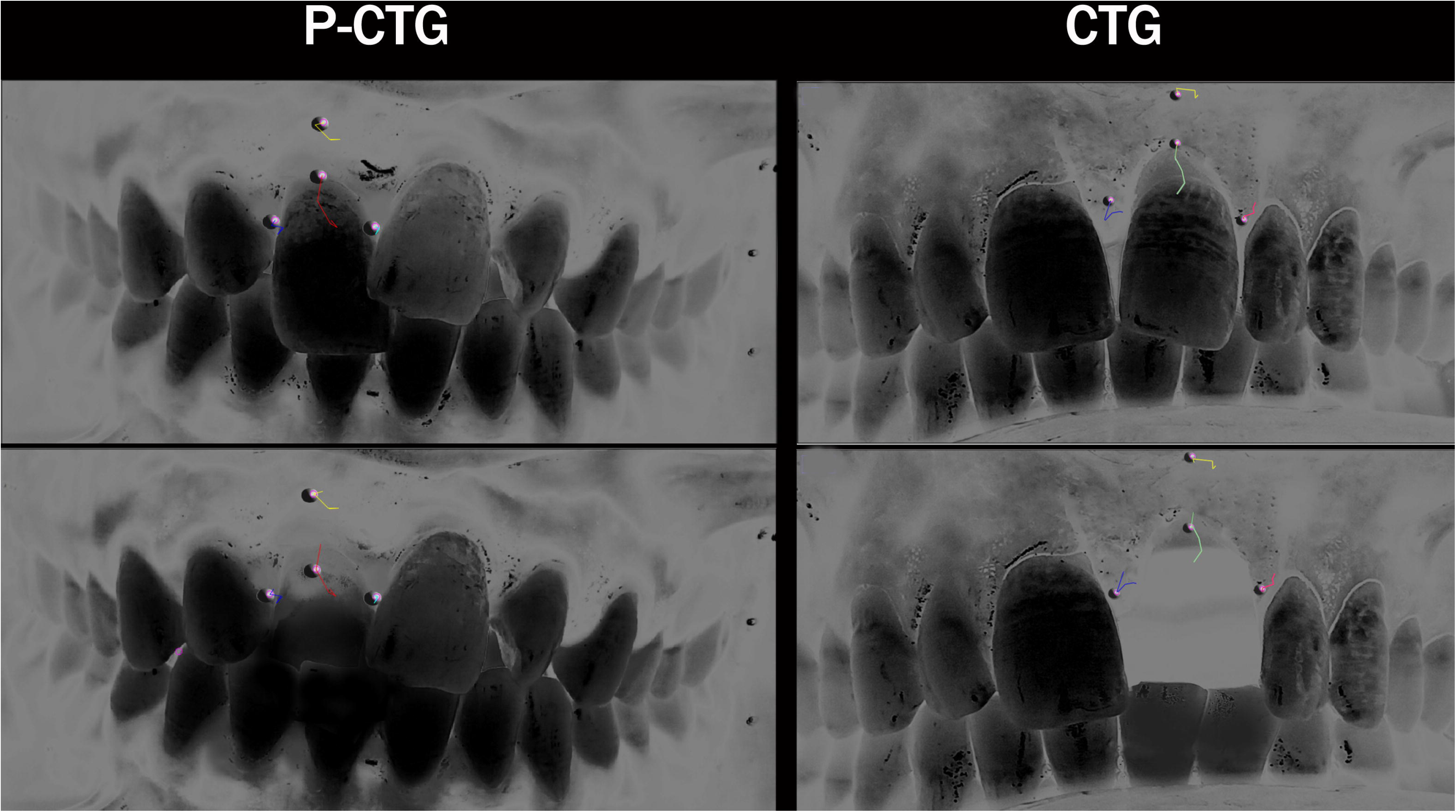
Tracks generated with TrackMate® plugin denoting the displacement of the four markers MG, MGJ, MP and DP in both the groups. Figure also shows the displacement of the markers before (top-left & right) and after extraction (bottom-left & right) after treatment with P-CTG and CTG respectively.

#### Bucco-lingual and mesiodistal dimensions of edentulous sites

The distance from the greatest convexity or bulge on the buccal and lingual/palatal soft-tissue beneath the buccal and lingual mucogingival lines (or the mid-buccal area immediately beneath the first rugae in the palatal mucosa [11,13,14]) at the center of the mesio-distal space was considered as the bucco-lingual dimension (Figure 4) [26]. This was measured by using a Vernier calipers at baseline, 3 and 6-months. The distance from the crests of the mesial to the distal papilla through the center of the ridge was considered as the mesio distal dimension [27]. A wet, 1 mm silk cord was adapted onto the ridge and marks were placed on the chord near the mesial and distal papilla. It was easy to adapt the silk cord on to the alveolar ridge with its irregularities than measuring it with a device. It was then retrieved and its length was measured with a Vernier calipers at baseline, 3 and 6-months (Figure 4). The difference from the baseline to 6-months in both the dimensions was calculated and was described as bucco-lingual gain (BL gain) and mesiodistal gain (ML gain) respectively.

### Statistical analysis

Data was analyzed with Statistical Package for Social Sciences (SPSS® v26.0, IBM Corp, Armonk, NY, USA). Friedman test and repeated measures ANOVA were used to compare data within the same group at different time intervals. Independent samples t-test and Mann-Whitney U test were used for comparing data between two groups. Pearson correlation coefficient was used to analyse linear relationships. Confidence intervals were set at 95% and *P* values ≤0.001 was considered as highly significant, *P*≤0.05 as significant and *P*>0.05 as not significant.

## RESULTS

All the participants (n=45; P-CTG, n=23; CTG n=22) completed study-related interventions. 5 participants (4 from P-CTG group and 1 from CTG group) withdrew after the intervention phase. Hence, statistical analysis was limited to 40 subjects (P-CTG, n=19; CTG n=21). A few adverse events such as swelling and minor bleeding episodes were conservatively managed in the post-operative healing phase.

### Displacement and total displacement of the four markers

There was a highly significant intragroup difference in displacement of MG in both the groups *(P=0.001)* and a significant intragroup difference in displacement of MGJ *(P=0.01)* and MP *(P=0.02)* in the CTG group between two time-points. No significant displacement of DP was seen in both the groups. *(P=0.09 & P=0.06* in P-CTG & CTG groups). Intergroup comparison of the displacement of markers across both the groups between two time-points showed varying levels of significance (Table 1). However, the total displacement of MG *(7.21 vs 6.11* mm) and MGJ *(4.59 vs 3.41* mm) was significantly higher in P-CTG group when compared to CTG group *(P=0.02)*. A highly significant displacement of MP *(P*≤*0.001)* was seen in CTG over the P-CTG group while no significant difference was seen in total displacement of DP in between the groups *(P=0.09)*.

**Table 1:** Comparison of the displacement (in *mm*) of the four markers, marginal gingiva (MG), mucogingival junction (MGJ), mesial papilla (MP) and distal papilla (DP), between two time-intervals across both the groups (P-CTG and CTG)

| Displacement (in mm) |  | Baseline -Post OP | Post OP- 1 month | 1 month- 3 months | 3 months- 6 months | Total Displacement (TD) (mm) | Intragroup Significance (F, P-value) |
| --- | --- | --- | --- | --- | --- | --- | --- |
| **MG** | **P-CTG** | 3.01±1.10 | 2.39±1.80 | 0.90±0.32 | 0.91±0.34 | 7.21±2.01 | 36, 0.001<sup>a)</sup><br>71, 0.001<sup>a)</sup> |
|  | **CTG** | 0.92±0.43 | 1.10±0.39 | 2.11±1.25 | 1.97±0.67 | 6.11±2.99 |  |
|  | **Intergroup Significance (t, p-value)** | 0.32, 0.001<sup>a)</sup> | 0.29, 0.001<sup>a)</sup> | 0.54, 0.001<sup>a)</sup> | 0.23, 0.001<sup>a)</sup> | 0.32, 0.02<sup>b)</sup> |  |
| **MGJ** | **P-CTG** | 0.79±0.13 | 1.99±0.76 | 0.92±0.44 | 0.89±0.66 | 4.59±1.70 | 165, 0.1<sup>c)</sup><br>54, 0.01<sup>b)</sup> |
|  | **CTG** | 0.92±0.28 | 1.80±0.40 | 0.47±0.29 | 0.22±0.05 | 3.41±1.83 |  |
|  | **Intergroup Significance (t, p-value)** | 0.96, 0.2<sup>c)</sup> | 0.89, 0.09<sup>c)</sup> | 0.92, 0.02<sup>b)</sup> | 0.89, 0.001<sup>a)</sup> | 0.81, 0.02<sup>b)</sup> |  |
| **MP** | **P-CTG** | 0.33±0.19 | 0.29±0.14 | 0.15±0.08 | 0.14±0.04 | 0.91±0.32 | 243, 0.9<sup>c)</sup><br>92, 0.02<sup>b)</sup> |
|  | **CTG** | 1.05±0.20 | 0.56±0.30 | 0.49±0.31 | 0.15±0.06 | 2.25±1.28 |  |
|  | **Intergroup Significance (t, p-value)** | 0.76, 0.001<sup>a)</sup> | 0.97, 0.01<sup>b)</sup> | 0.54, 0.001<sup>a)</sup> | 0.2, 0.9<sup>c)</sup> | 0.66, 0.001<sup>a)</sup> |  |
| **DP** | **P-CTG** | 0.46±0.31 | 0.55±0.29 | 0.59±0.22 | 0.36±0.21 | 1.96±0.91 | 146, 0.09<sup>c)</sup><br>29, 0.06<sup>c)</sup> |
|  | **CTG** | 0.22±0.07 | 0.54±0.20 | 0.42±0.16 | 0.52±0.23 | 2.46±1.73 |  |
|  | **Intergroup Significance (t, p-value)** | 0.27, 0.01<sup>b)</sup> | 0.92, 0.9<sup>c)</sup> | 0.45, 0.1<sup>c)</sup> | 0.68, 0.01<sup>b)</sup> | 0.76, 0.09<sup>c)</sup> |  |

<sup>a)</sup> Highly significant <sup>b)</sup> Significant <sup>c)</sup> Non-significant CTG: Connective tissue graft group, P-CTG: Pediculated connective tissue graft group, MG: Marginal gingiva, MGJ: Mucogingival junction, MP: Mesial papilla, DP: Distal papilla, TD: Total displacement.

### Divergence and angle of maximum divergence of the four markers

Intragroup analysis revealed a highly significant difference in divergence of MG in both the groups *(P*≤*0.001)* and significant difference in the divergence of MGJ *(P-CTG P=0.03, CTG P=0.04)*, MP *(P-CTG P=0.05, CTG P=0.04)* and DP *(P-CTG P=0.05, CTG P=0.05)* at various time intervals. Varying levels of significance were seen in the intergroup comparisons of divergence across both the groups between two time-points (Table 2). The AMD was significantly higher in CTG group for MG *(P=0.01)* and MGJ *(P=0.01)* when compared to P-CTG group. For MP and DP, a highly significant *(P*≤*0.001)* AMD was seen in CTG and P-CTG group respectively.

**Table 2:**
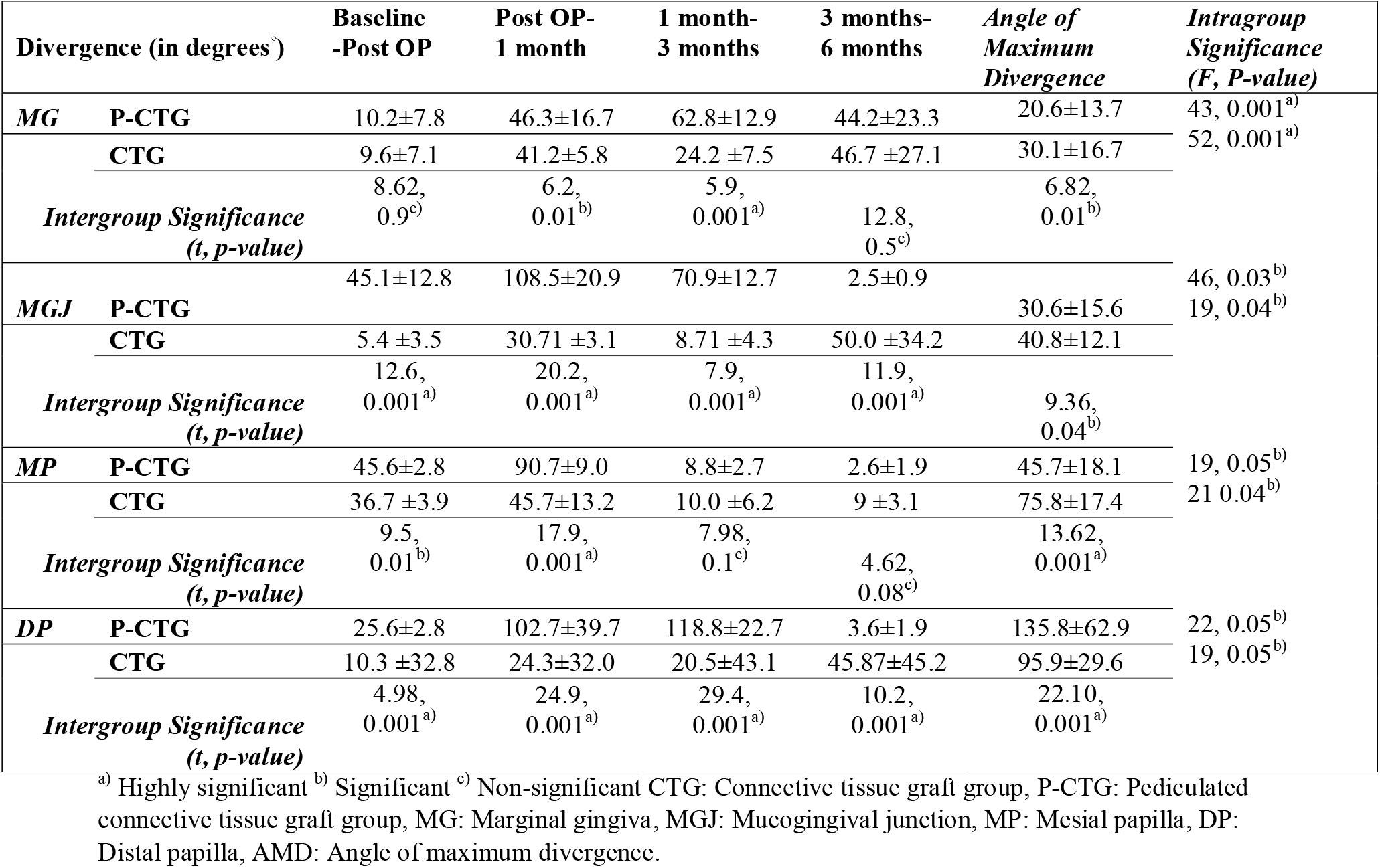
Comparison of divergence and angle of maximum divergence (in degrees°) of the four markers, marginal gingiva (MG), mucogingival junction (MGJ), mesial papilla (MP) and distal papilla (DP), between two time-intervals across both the groups (P-CTG and CTG)

| Divergence (in degrees°) |  | Baseline<br>-Post OP | Post OP-1 month | 1 month-3 months | 3 months-6 months | Angle of<br>Maximum<br>Divergence | Intragroup<br>Significance<br>(F, P-value) |
| --- | --- | --- | --- | --- | --- | --- | --- |
| **MG** | **P-CTG** | 10.2±7.8 | 46.3±16.7 | 62.8±12.9 | 44.2±23.3 | 20.6±13.7 | 43, 0.001<sup>a)</sup><br>52, 0.001<sup>a)</sup> |
|  | **CTG** | 9.6±7.1 | 41.2±5.8 | 24.2 ±7.5 | 46.7 ±27.1 | 30.1±16.7 |  |
|  | **Intergroup Significance**<br>**(t, p-value)** | 8.62,<br>0.9<sup>c)</sup> | 6.2,<br>0.01<sup>b)</sup> | 5.9,<br>0.001<sup>a)</sup> | 12.8,<br>0.5<sup>c)</sup> | 6.82,<br>0.01<sup>b)</sup> |  |
| **MGJ** | **P-CTG** | 45.1±12.8 | 108.5±20.9 | 70.9±12.7 | 2.5±0.9 | 30.6±15.6 | 46, 0.03<sup>b)</sup><br>19, 0.04<sup>b)</sup> |
|  | **CTG** | 5.4 ±3.5 | 30.71 ±3.1 | 8.71 ±4.3 | 50.0 ±34.2 | 40.8±12.1 |  |
|  | **Intergroup Significance**<br>**(t, p-value)** | 12.6,<br>0.001<sup>a)</sup> | 20.2,<br>0.001<sup>a)</sup> | 7.9,<br>0.001<sup>a)</sup> | 11.9,<br>0.001<sup>a)</sup> | 9.36,<br>0.04<sup>b)</sup> |  |
| **MP** | **P-CTG** | 45.6±2.8 | 90.7±9.0 | 8.8±2.7 | 2.6±1.9 | 45.7±18.1 | 19, 0.05<sup>b)</sup><br>21 0.04<sup>b)</sup> |
|  | **CTG** | 36.7 ±3.9 | 45.7±13.2 | 10.0 ±6.2 | 9 ±3.1 | 75.8±17.4 |  |
|  | **Intergroup Significance**<br>**(t, p-value)** | 9.5,<br>0.01<sup>b)</sup> | 17.9,<br>0.001<sup>a)</sup> | 7.98,<br>0.1<sup>c)</sup> | 4.62,<br>0.08<sup>c)</sup> | 13.62,<br>0.001<sup>a)</sup> |  |
| **DP** | **P-CTG** | 25.6±2.8 | 102.7±39.7 | 118.8±22.7 | 3.6±1.9 | 135.8±62.9 | 22, 0.05<sup>b)</sup><br>19, 0.05<sup>b)</sup> |
|  | **CTG** | 10.3 ±32.8 | 24.3±32.0 | 20.5±43.1 | 45.87±45.2 | 95.9±29.6 |  |
|  | **Intergroup Significance**<br>**(t, p-value)** | 4.98,<br>0.001<sup>a)</sup> | 24.9,<br>0.001<sup>a)</sup> | 29.4,<br>0.001<sup>a)</sup> | 10.2,<br>0.001<sup>a)</sup> | 22.10,<br>0.001<sup>a)</sup> |  |

<sup>a)</sup> Highly significant <sup>b)</sup> Significant <sup>c)</sup> Non-significant CTG: Connective tissue graft group, P-CTG: Pediculated connective tissue graft group, MG: Marginal gingiva, MGJ: Mucogingival junction, MP: Mesial papilla, DP: Distal papilla, AMD: Angle of maximum divergence.

### Bucco-lingual and mesiodistal dimensions of edentulous sites

No significant intergroup difference was observed between the P-CTG and CTG groups for buccolingual dimensions at baseline *(P=0.601)*. There was a statistically higher increase in the buccolingual dimensions in the P-CTG group at 3 months *(P=0.001)* and 6 months *(P=0.002)* when compared to the CTG group. There was a gain (BL gain) of 2.82±1.29 mm and 1.68±1.12 mm from baseline to 6-months in P-CTG and CTG group respectively. No significant difference was observed between P-CTG and CTG groups for the mesiodistal dimensions of the edentulous sites at baseline, 3-months and 6-months (*P=0.482, P=0.459* & *P=0.561*). There was a gain (MD gain) of 1.27±0.65 mm and 1.48±0.67 mm from baseline to 6-months in P-CTG and CTG group respectively.

### Correlation between TD and AMD of the four markers *vs* MD and BL gain

Correlation analysis of AMD for MG and BL gain revealed a moderate positive correlation MG *(r=0.5, P=0.05)* in P-CTG group. In CTG group, strong *(r=0.7, P=0.03)* and a moderate positive correlation *(r=0.3, P=0.04)* between AMD for MGJ and AMD for MP with BL gain was seen. A moderate positive correlation was seen *(r=0.4, P=0.001)* between TD of MG and BL gain in P-CTG group. A weak negative correlation was seen *(r=-0.2, P=0.005)* between TD of DP and MD gain in CTG group. Weak positive correlation *(r=0.2, P=0.05)* was seen between TD of MGJ and BL gain in P-CTG group and strong positive correlation was seen between TD of MP and BL gain in CTG group *(r=0.7, P=0.04)* (Table 3).

**Table 3:** Correlation between TD and AMD of the four markers *vs* MD and BL gain across both the groups (P-CTG and CTG)

| Correlations |  | TD vs BL Gain |  | TD vs MD Gain |  | AMD vs BL Gain |  | AMD vs MD Gain |  |
| --- | --- | --- | --- | --- | --- | --- | --- | --- | --- |
| Pearson correlation |  | *r* | *P* | *r* | *P* | *r* | *P* | *r* | *P* |
| **MG** | **P-CTG** | 0.4 | 0.001<sup>a)</sup> | -0.2 | 0.21 | 0.5 | 0.05<sup>b)</sup> | -0.3 | 0.18 |
|  | **CTG** | 0.8 | 0.06 | 0.4 | 0.89 | -0.1 | 0.21 | 0.1 | 0.98 |
| **MGJ** | **P-CTG** | 0.2 | 0.05<sup>b)</sup> | -0.4 | 0.12 | 0.2 | 0.39 | 0.61 | 0.19 |
|  | **CTG** | 0.1 | 0.15 | 0.1 | 0.13 | 0.7 | 0.03<sup>b)</sup> | 0.1 | 0.2 |
| **MP** | **P-CTG** | 0.1 | 0.45 | -0.1 | 0.36 | 0.3 | 0.1 | 0.4 | 0.33 |
|  | **CTG** | 0.7 | 0.04<sup>b)</sup> | 0.2 | 0.13 | 0.3 | 0.04<sup>b)</sup> | -0.2 | 0.11 |
| **DP** | **P-CTG** | 0.1 | 0.09 | 0.1 | 0.18 | 0.1 | 0.12 | 0.1 | 0.07 |
|  | **CTG** | 0.1 | 0.12 | -0.2 | 0.05<sup>b)</sup> | 0.1 | 0.06 | 0.1 | 0.2 |

a) Highly significant b) Significant *r* = Pearson correlation coefficient
CTG: Connective tissue graft group, P-CTG: Pediculated connective tissue graft group, MG: Marginal gingiva, MGJ: Mucogingival junction, MP: Mesial papilla, DP: Distal papilla, TD: Total displacement, AMD: Angle of maximum divergence, BL gain: Buccolingual gain, ML gain: mesiodistal gain.

## DISCUSSION

There exist studies that have sought to locate and analyse alterations in specific landmarks before and after mucogingival procedures [29-31]. Periodontal probing under magnification [30] or with a surgical stent [31], and techniques such as reproducible photography [22] and radiographic soft tissue determination have been used previously [15,22]. In this study we have used a software system based on the principles of image alignment [22], stacking [23] and tracking [24,25] to capture movement of specific landmarks before and after soft tissue grafting. Though designed initially for multidimensional microscopy, TrackMate® can track movements in millimetres *(mm)*[28], is flexible and various researchers, including periodontists, can quickly develop an algorithm suited for specific projects.

Both the treatment modalities resulted in a significant displacement and divergence of the markers; Relative to each other, P-CTG resulted in a significant displacement of the mucogingival junction and marginal gingiva; whereas CTG resulted in a marked divergence of all the markers except distal papilla. The use of P-CTG and CTG in general induces a robust inflammatory and healing response that initially results in an oedema and later results in an increased graft density and thickening of the epithelium which may have resulted in the movement of these markers [12,26,29].

The displacement of marginal gingiva by P-CTG was greater than CTG (7.21 vs 6.11 mm); Two reasons can be ascribed to this finding; 1. Being a pediculated graft, the quality of healing is much higher and the chance of formation of a thick biotype is greater resulting in an increased soft tissue fill and subsequent displacement of landmarks [10,12,14,17,26,29] and 2. There is negligible disturbance of the marginal gingiva as there was a minimal elevation of the buccal flap[13,14]. In CTG, elevation of the flap to ensure adaptation to the recipient site is necessary and the volume of the graft itself may result in the post-operative margin being apical to the pre-operative levels [7,8,10]. The integrity and level of this postoperative gingival margin is crucial [14,19] as it seems to augment coronal migration during the subsequent healing phase [30]. The two reasons may also explain the greater displacement of MGJ by P-CTG over CTG (4.59 vs 3.41 mm). We also observed a mean immediate post-surgical MGJ shift of 0.79 mm and 0.92 mm for P-CTG and CTG respectively. These are lower than values obtained for other procedures. Pini Prato [30] and Saber [31] reported a post□surgical shift of 4.03 mm and 2.11 mm for coronally advanced flap and subepithelial connective tissue graft respectively. In both these procedures, there was a substantial flap advancement [3,4,8] which may have contributed to greater shift of MGJ.

The manipulation of buccal tissues also resulted in a significant deviation of all markers from their original positions (except DP) in the CTG group. Previous studies [30-32] have demonstrated that MGJ seems to shift between 0.8 to 2 mm coronally or apically within 6-months. Rather than a linear drop, we observed a semi-angle to horizontal movement of the MGJ from baseline to the end-of-study period in P-CTG and CTG groups respectively. A movement of MGJ of 3-4 mm at an angle of 30° to 40° may affect aesthetic or functional outcomes; Nemcovsky & Artzi [33], Goldstein et al. [34] and Bontá [9] have advocated a palatal approach to achieve socket closure primarily to prevent alterations in MGJ position. Their observations correspond with the lesser degree of divergence seen with P-CTG in our study. With either the CEJ [31,32] or the incisal margin [30] as a reference point, previous studies have stated that the average apical shift of MGJ reduces to around 1mm from 6-months onwards [30,32]; Probing or measuring a distance between two landmarks is along a single axis/direction and may not detect deviations in other planes or directions[15,23,24,25]. The plugin used in this study can track a marker in two axes; We were able to observe a more angulated to horizontal movement of the MGJ in both the groups (Figure 5). Both Saber [31] and Gürgan et al. [32] in their respective studies on subepithelial connective tissue graft and coronally repositioned flap observed that the MGJ reverts to its original position between 1 to 5 years. Our study duration was shorter and it is not possible to compare results with the aforementioned studies.

Mixed results were seen across the two groups with regard to the movement of the papillary markers MP and DP. CTG resulted in a significant movement of MP & DP and divergence of the marker MP over P-CTG. The P-CTG surgical protocol involves placing the pedicle passively over the extraction site with minimal manipulation of the papilla and *vice versa* [10,13,14]. CTG results in an increase in soft tissue volume in the papillary region [7,10,11] resulting in the displacement and deviation of these markers. Interestingly, correlation analysis revealed that the insignificant displacement of the marker DP resulted in an increased soft tissue gain in the mesiodistal dimension. Elevation of the flap for CTG placement results in a lower papilla height [16,35]; in one study however, lower height of papillae was actually associated with better clinical outcomes [36].

In the P-CTG group, correlation analysis revealed that the TD and maximum divergence (AMD) of MG resulted in a significant gain of soft tissue in the buccolingual dimension (moderately strong correlation between TD & AMD of MG and BL gain). Though the correlation was weak, total displacement and maximum divergence of MGJ resulted in BL gain as well. P-CTG also resulted in higher buccolingual dimensions at 3 months *(P=0.001)* and 6 months when compared to the CTG group. The surgical protocol itself [13,14], thicker pedicle and epithelial keratinization [12,26,29], formation of a thick biotype [10,12,14,17], the pre-operative integrity of the gingival margin [14,19] and substantial flap advancement [3,4,8] are probably the factors contributing to this gain in soft tissue. In the CTG group, total displacement and maximum deviation of MP and maximum deviation of MGJ resulted in a significant gain of soft tissue in the buccolingual dimension. Increase in soft tissue volume in the papillary region [7,8] a lower but stable papilla height [16,35], horizontal rather than apico-coronal movement of the MGJ which leads to an increase in width of keratinised tissue [29-31], flap advancement [3,4] and its general predictability of volume gain [2] can be the reasons for this positive effect.

No significant difference was observed between P-CTG and CTG groups for mesiodistal dimensions at baseline, 3-months and 6-months. While there was a mesiodistal gain (MD gain) of 1.27 and 1.48 mm from baseline to 6-months in the P-CTG and CTG group respectively, the displacement and divergence of the markers seem to have no effect on this outcome. The mesio-distal dimensions are determined by the papillary heights, which in turn are dependent on the integrity of the crests of the bone in this region [16,27]. In their studies on hard-tissue augmentation in extraction sockets, Barone et al [37] and Aimetti et al [38], noted a slight reduction in mesiodistal dimensions by 0.4-0.5 mm at 3 to 7 months despite grafting the sockets with bone-grafts. Considering that no hard-tissue grafting has been done in the participants, a similar effect can be expected in this study as well [5,7].

This study has some limitations worth noting. While a combined hard and soft-tissue grafting would have yielded better outcomes [3,9,17,26], hard-tissue grafting was not done in the patient’s best interest. This was also the reason why we did not evaluate radiographic landmarks in this study. The authors are not aware of any clinical trials in dentistry which have used the TrackMate® plugin; hence a new protocol and algorithm for tracking markers was developed by the authors themselves. The terms ‘divergence’ and ‘angle of maximum divergence’ are not specific terms or definitions; rather they are generic adaptations of terms from key articles [23-25,28] developed for better understanding of the outcomes.

To conclude, this study evaluated the effect of connective tissue grafts (CTG) and pediculated connective tissue grafts (P-CTG) for socket closure on soft tissue landmarks and ridge dimensions. P-CTG and CTG had significant effects on displacement and divergence of soft tissue landmarks respectively. The relocation of these landmarks seems to result in an improvement in the buccolingual dimensions of the post-extraction ridge.

## Data Availability

All data produced in the present work are contained in the manuscript

## ACKNOWLEDGEMENTS

The study was wholly supported by the author’s institutions and received no external support.

